# Exploring autism and autism co-occurring condition associations to elucidate multivariate genetic mechanisms and insights

**DOI:** 10.1101/2024.01.07.24300940

**Authors:** Karoliina Salenius, Niina Väljä, Sini Thusberg, Francois Iris, Christine Ladd-Acosta, Christophe Roos, Matti Nykter, Alessio Fasano, Reija Autio, Jake Lin, the GEMMA study

**Affiliations:** Faculty of Medicine and Health Technology, Tampere University and Tays Cancer Centre, Tampere, Finland; The GEMMA study, GEMMA-PROJECT.eu, Salerno, Italy; BMSystems, Paris, France; Department of Epidemiology, Johns Hopkins Bloomberg School of Public Health, Baltimore, USA; Euformatics, Tekniikantie, Espoo, Finland; Foundation for the Finnish Cancer Institute, Helsinki, Finland; European Biomedical Research Institute of Salerno (EBRIS), Salerno, Italy; Harvard Medical School, Harvard T.H. Chan School of Public Health, Boston, USA; Health Sciences, Faculty of Social Sciences, Tampere University, Tampere, Finland; Department of Medical Epidemiology and Biostatistics, Karolinska Institute, Stockholm, Sweden

**Keywords:** Autism biomarkers, Multivariate GWAS, Mendelian randomization, GEMMA

## Abstract

**Background:** Autism is a partially heritable neurodevelopmental condition, and people with autism may also have other co-occurring conditions such as ADHD, anxiety disorders, depression, mental health issues, learning difficulty, physical health conditions and communication challenges. The concomitant development of autism and other neurological conditions is assumed to result from a complex interplay between genetics and the environment. However, only a limited number of studies have performed analysis on multivariate genetic autism associations.

**Methods:** We conducted to-date the largest multivariate GWAS on autism and 8 autism co-occurring condition traits (ADHD, ADHD childhood, anxiety stress, bipolar, disruptive behaviour, educational attainment, major depression, and schizophrenia) using summary statistics from leading studies. Multivariate associations and central traits were further identified. Subsequently, colocalization and Mendelian randomization (MR) analysis were performed on the associations identified with the central traits containing autism. To further validate our findings, pathway and quantified trait loci (QTL) resources as well as independent datasets consisting of 92 (30 probands) whole genome sequence data from the GEMMA project were utilized.

**Results:** Multivariate GWAS resulted in 637 significant associations (p < 5e-8), among which 322 are reported for the first time for any trait. 37 SNPs were identified to contain autism and one or more traits in their central trait set, including variants mapped to known SFARI autism genes MAPT and NEGR1 as well as novel ASD genes KANSL1, NSF and NTM, associated with immune response, synaptic transmission, and neurite growth respectively. Mendelian randomization analyses found that all 8 co-occuring conditions are associated with autism while colocalization provided strong evidence of shared genetic aetiology between autism and education attainment, schizophrenia and bipolar traits. Allele proportions differences between MAPT (17q21.31) region aberrations and MAPT H1/H2 haplotypes, known to associate with neurodevelopment wwere found between GEMMA autism probands and controls. Pathway, QTL and cell type enrichment implicated microbiome, enteric inflammation, and central nervous system enrichments.

**Conclusions:** Our study, combining multivariate genome-wide association testing with systematic decomposition identified novel genetic associations related to autism and autism co-occurring driver traits. Statistical tests were applied to discern evidence for shared and interpretable liability between autism and co-occurring traits. These findings expand upon the current understanding of the complex genetics regulating autism and reveal insights of neuronal brain disruptions potentially driving development and manifestation.

**Highlights:**

- Multivariate GWAS resulted in 637 significant ASD associations (p < 5e-8), among which 322 are reported for the first time.
- The novel associations mapped to known SFARI autism genes MAPT and NEGR1 and novel ASD markers KANSL1, NSF and NTM markers, associated with immune response, synaptic transmission, and neurite growth, potentially driving the gut brain-barrier hypothesis driving ASD.
- Mendelian randomization analyses found that the co-occuring traits ADHD, ADHD childhood, anxiety stress, bipolar, disruptive behaviour, educational attainment, major depression, and schizophrenia are strongly associated with autism.

## Introduction

Autism spectrum disorders (autism) is an umbrella term for a group of heterogeneous neurodevelopmental conditions that manifest in early childhood. Autism is associated with polygenic markers and with several key factors such as family history, genetics and environment (Khachadourian et al. 2023; Nayar et al. 2021; Chaste and Leboyer 2012). The diagnosis of autism is based on its key characteristics including difficulties in social communication and interaction, restricted and repetitive behaviors, hyperactivity and divergent responses to sensory inputs. The most common co-occurring conditions in autistic persons are attention deficit hyperactivity disorder (ADHD), ADHD childhood, anxiety, bipolar (BP), depression, epilepsy, obsessive compulsive disorders (OCD) and stress related conditions, all of which share overlapping diagnostic attributes and challenging symptoms with autism (Khachadourian et al. 2023; Romero et al. 2016). According to US data, autistic children tend to fare less well in educational attainment (EA) and about one in three have a reduced intellectual ability, as defined by intelligence quotient (IQ less than 70) (Baio et al. 2018; Tamm et al. 2020). Some children with autism having higher IQ scores also comparatively experience harder academic struggles due to co-occurring conditions and difficulties in social interactions (Ashburner, Ziviani, and Rodger 2010).

Together with recent advances in genomics technology and pivotal support from the engaged autism community, 1,162 genes are currently implicated with autism development and these are curated in the SFARI (Arpi and Simpson 2022; Grove et al. 2019; Pinto et al. 2014) gene module. These genes, with varying degrees of effect, are scored using the Evaluation of Autism Gene Link Evidence (EAGLE) framework (Schaaf et al. 2020). Surprisingly, while it is known that common variants contribute to the majority of genetic background (Gaugler et al. 2014), only a few robust genetic associations have been recently reported. Most of these are attributed to the landmark study conducted by Grove and colleagues, employing a large Danish cohort with 18,381 autism cases and 27,969 controls, where 12 significant variant associations were reported (Grove et al. 2019).

Given that there is overlap in symptoms between autism and ADHD, a genetics study recently found shared genetic factors underlying autism and ADHD (Peyre et al. 2021), with partial concordance between bidirectional colocalization single nucleotide variants (SNPs). However, the study was limited to general ADHD (onset age 10+), and not childhood ADHD. Astoundingly many (47% median) autistic children have reported one or more gastrointestinal (GI) symptoms (Holingue et al. 2018; Boorstein 2008). Recently, there have been promising results that link microbiome disruption and diversity (Morton et al. 2023) as a novel contributing factor to autism. Interestingly, while Grove and colleagues found that 7 of the 12 autism SNP associations have similar significance towards EA and psychosis traits depression and schizophrenia (Grove et al. 2019), still very little is known concerning the joint and the shared genetic mechanisms between autism and autism co-occurring traits including ADHD, ADHD childhood, EA, depression and their potential links to gastrointestinal disruptions.

To attenuate on the genetic knowledge gaps in autism and expand the exploration of potential shared co-occurring trait genetic associations, this study performed multivariate genome wide association with summary statistics from autism and 12 co-occurring traits from large reputable cohorts. To achieve this, colocalization (coloc) was systematically applied to test the robustness between the shared variants and traits (Wallace 2021). Mendelian randomisation (MR) was further applied, using the multivariate variants and the essential traits, to assess liability relationships between autism and the selected co-occurring conditions (Bowden, Davey Smith, and Burgess 2015; Rees, Wood, and Burgess 2017). This study seeks to clarify functional, regulatory and tissue type differentiation with enrichment and integration of quantified trait loci (QTL) integration while validating our key findings with independently sequenced genomes from the GEMMA cohort (Troisi et al. 2020).

## Methods and Materials

Genome-wide summary statistics for autism and ADHD were collected from the Psychiatric Genomics Consortium (PGC) and iPSYCH (Pedersen et al. 2018; Sullivan et al. 2018) studies. Education attainment (Okbay et al. 2022) summary file was collected from the Social Science Genetic Association Consortium (SSGAC). Additional autism co-occurring traits, selected based on LDSC genetic correlation with autism, include ADHD childhood, bipolar (BP), anxiety-stres disorder (ASRD), disruptive behaviour (DBD), major depression (MDD) and schizophrenia (SCZ), with sample sizes ranging from 31,890-765,283 are shown in Table 1 (additional details in Supplementary Table 1). Summary statistics are joined, yielding 4,525,476 SNPs, and applied in a multivariate GWAS setting. Follow-up analysis includes decomposition aiming to detect th most important traits while colocalization and Mendelian randomisation analysis are conducted to explore shared liability as shown in Figure 1.

**Figure 1:**
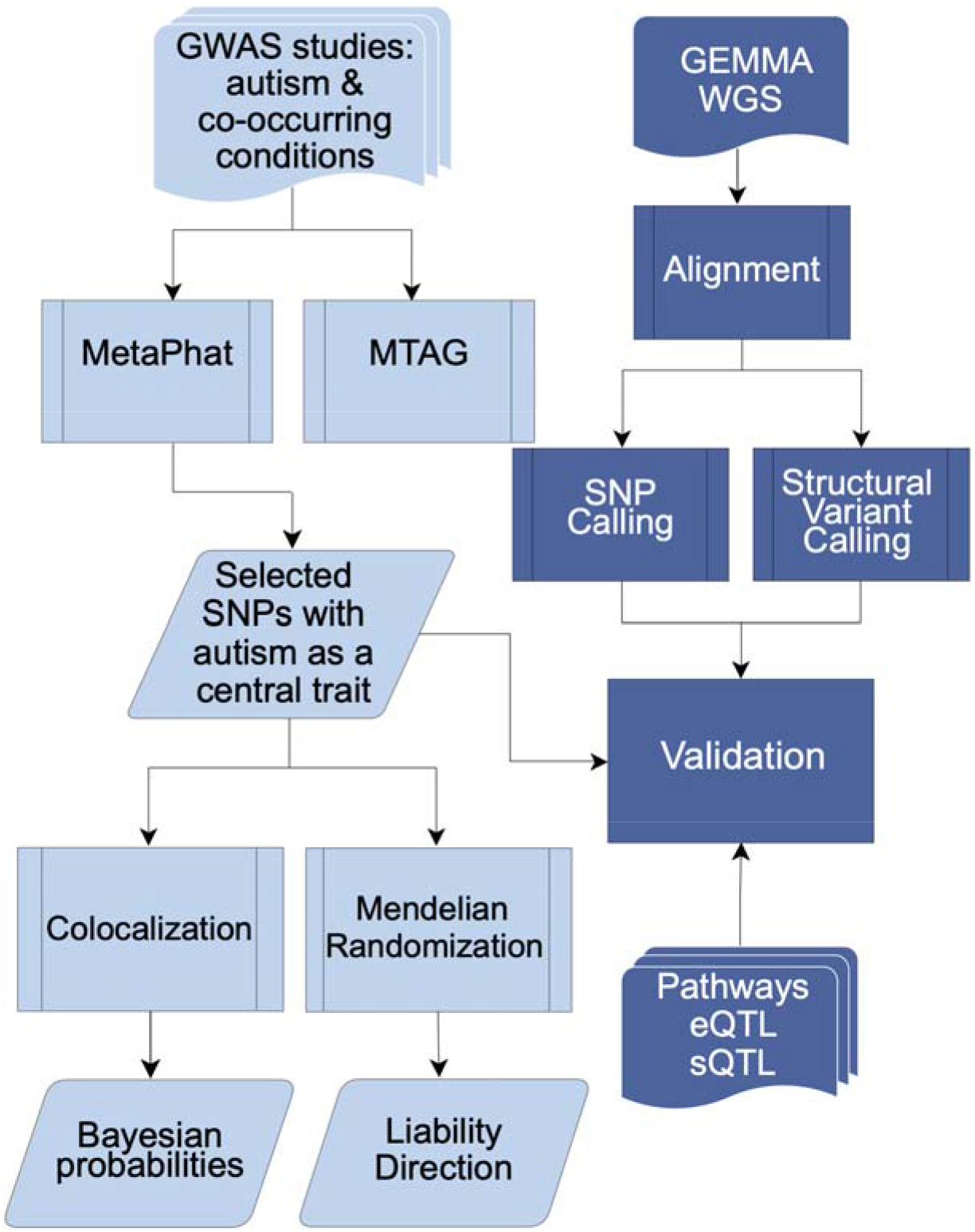
Workflow for the analyses conducted in the study. Multivariate GWAS was performed on selected GWAS studies including autism and 8 co-occurring traits: ADHD, ADHD childhood, bipolar, anxiety, disruptive behaviour, educational attainment, major depression and schizophrenia. 37 SNPs were selected and evaluated with Colocalization and Mendelian Randomization. Further validation of these SNPs utilized pathway and EBI eQTL/sQTL catalogs as well as the GEMMA -study. The GEMMA whole genome sequencing (WGS) processing included variant calling to infer structural and single nucleotide variants (SVs and SNVs) present in the samples.

**Table 1.** Data of autism and 8 genetically correlated traits (P < 0.05, calculated from LDSC) are presented and applied towards multivariate-GWAS to explore multivariate associations and additional trait refinement. More details and excluded traits are listed in Supplementary Table 1.

| **Trait** | **Heritability (H2)** | **Genetic correlation** | **P** | **Genetic covariance** | **Intercept** |
| --- | --- | --- | --- | --- | --- |
| Autism<sup>1</sup> | 0.203 (0.015) | na | na | na | na |
| ADHD<sup>2</sup> | 0.094 (0.005) | 0.535 (0.041) | 1,44E-38 | 0.074(0.006) | 0.233 (0.009) |
| ADHD Childhood<sup>3</sup> | 0.235 (0.015) | 0.478 (0.052) | 5,21E-20 | 0.104 (0.012) | 0.260 (0.007) |
| ASRD<sup>4</sup> | 0.211 (0.019) | 0.441 (0.079) | 2,22E-08 | 0.090 (0.015) | 0.221 (0.006) |
| Bipolar<sup>5</sup> | 0.068 (0.003) | 0.219 (0.041) | 9,67E-08 | 0.026 (0.005) | 0.032 (0.006) |
| DBD<sup>6</sup> | 0.1004 (0.012) | 0.186 (0.07) | 0,008 | 0.026 (0.010) | 0.179 (0.006) |
| EA<sup>7</sup> | 0.321 (0.009) | 0.207 (0.025) | 9,95E-17 | 0.053 (0.007) | -0.005 (0.007) |
| MDD<sup>8</sup> | 0.026 (0.002) | 0.505 (0.003) | 2,78E-36 | 0.037 (0.003) | 0.155 (0.005) |
| SCZ<sup>9</sup> | 0.395 (0.014) | 0.258 (0.035) | 7,87E-14 | 0.07 (0.01) | 0.018 (0.007) |

### Multivariate GWAS and determination of central traits

Multivariate GWAS on autism and autism co-occurring biomarker traits were performed using metaPhat/metaCCA software that performs multivariate analysis by implementing Canonical Correlation Analysis (CCA) for a set of univariate GWAS summary statistics (Ruotsalainen et al. 2021; Cichonska et al. 2016; Lin et al. 2020). The objective of metaCCA is to find the optimal genetic effect combination that is maximally correlated with a linear combination of the trait variables. Autism multivariate central traits are identified by MetaPhat decomposition based on iterative tracing of p-values (p) from trait subsets (relative to 5×10-8) and Bayesian Information Criterion (BIC) (Schwarz 1978) representing model fit. The central traits are the union of the driver biomarker traits together with the subset traits yielding the lowest BIC value. MTAG (Turley et al. 2018), a high performant multivariate-GWAS that addresses sample overlap, is additionally performed for validation.

### Genetic annotations, pathway enrichment and validation

SFARI Base Gene resource, GeneCards and GWAS catalog were used to assess the novelty of variants and genes associated with autism (Arpi and Simpson 2022; Safran et al. 2010; MacArthur et al. 2017). snpXplorer was applied towards SNP annotation(Tesi et al. 2021). Reactome and WikiPathway databases pathway enrichments were evaluated with the Enrichr tool (Kuleshov et al. 2016). Human organ and cell type systems enrichment analysis, encompassing 1,466 tissue-cell type and single-cell RNAseq panels, was conducted using WebCSEA (Dai et al. 2022, Lake et al. 2018). eQTL and sQTL were assessed within the QTL catalog, via FIVEx portal (Kwong et al. 2022).

### Colocalization analyses

Colocalization was performed for the selected multivariate autism SNPs to assess if the associated variants in the locus are shared genetically between autism and the 8 co-occurring related autism traits to account for erroneous results that may follow from analyzing individual SNPs. Errors can occur when a SNP associated with trait 1 and trait 2 are in linkage disequilibrium (LD). The analyses were performed using the R package coloc (Giambartolomei et al. 2014; Wallace 2020).

The colocalization analysis was conducted using the absolute base factor colocalization method (coloc.abf), which is a Bayesian colocalization analysis method. A region size window of 100KB (50±KB flanking the SNP position) was selected to comprehensively span potential LD and regulatory elements (Piovesan et al. 2016). The different hypotheses tested include: H0 (no liable variant), H1 (liable variant only for trait 1), H2 (liable variant only for trait 2), H3 (two separate liable variants), H4 (common liable variant shared between the traits). As recommended (Wallace 2020), default setting prior probability thresholds were applied: 1e-4 for H1, H2 and H3 and 1e-5 for H4 while posterior probability (H4 > 0.90) is conservatively applied to estimate shared liability.

### Mendelian randomization analyses

Mendelian Randomization analyses (MR) was conducted on the selected autism multi-trait SNPs based on their assigned central traits, to explore the liability, direction and independent (reverse causation) relationships between autism and its related biomarkers (Phillips and Smith 1991). Instrumental strengths, approximated with F1 score > 10, were calculated using SNP effect and standard error values (Bound, Jaeger, and Baker 1995; Palmer et al. 2012). The analyses were performed using the R package MendelianRandomization (Bowden, Davey Smith, and Burgess 2015).

### Whole genome sequencing

The results were validated using yet unpublished data from the EU Horizon2020 GEMMA research project with genotype variant calls in 97 (30 proband) WGS samples from the GEMMA prospective cohort (Troisi et al. 2020). 49 of these samples were sourced from 15 Italian families, 19 samples from 6 US families and 25 samples from 7 Irish families. These samples, assayed on whole blood and collected during enrollment, were sequenced with 30-40X coverage on Illumina NovaSeq 6000 platform. Data was aligned to GRCh38 reference genome using bwa mem v0.7.17 (Li 2013) and reads were sorted and duplicates marked with samtools v1.12 (Li et al. 2009). Quality control was performed with omnomicsQ -software (Gutowska-Ding et al. 2020). For variant calling DeepVariant v1.4.0 (Poplin et al. 2018) was utilized and variants were annotated with Variant Effect Predictor (McLaren et al. 2016) version 111.0.. Presence of the H1H2 inversion was evaluated based on haplotype defining variant rs8070723 (Allen et al. 2014). Structural variants were called with Smoove v0.2.6, a wrapper for Lumpy v0.3.1. to detect the 238bp deletion in MAPT gene intron 9 and to further validate the presence of the 900kb H1H2 inversion (Pittman, Fung, and de Silva 2006), which is not reliably detected from short read sequencing.

### Statistical analysis

All statistical analyses were performed using R 4.2.2 software and available as R markdown results in the github project (https://github.com/jakelin212/mvasd_gwas). Genome-wide association is called on the standard and strict p-value threshold of 5e-8, to account for multiple testing based on the assumption of about 1-million independent tests (Risch and Merikangas 1996). Bonferroni correction is applied for multiple testing.

## Results

### GWAS summary statistics

Genome-wide summary statistics for autism and ADHD were collected from the PGC and iPSYCH (Pedersen et al. 2018; Sullivan et al. 2018) studies. Education attainment (Okbay et al. 2022) summary file was collected from the Social Science Genetic Association Consortium (SSGAC). Altogether, using summary statistics, 12 autism co-occurring traits were assessed for genetic correlation with the landmark autism study (Grove et al. 2019), the largest genetic correlation values, as computed by LDSC (Bulik-Sullivan et al. 2015), were between autism and ADHD (0.535), followed by MDD (0.505) and ADHD childhood (0.478). Shown in Table 1 below, 8 traits are shown to be genetically correlated with autism (p < 0.05) and additional details of all traits are shown in Supplementary Table 1.

### Multivariate autism central trait SNPs, pathway and organ tissue enrichment

Multivariate GWAS was performed with autism together with its genetically correlated traits, ADHD, ADHD childhood, ASRD, bipolar, DBD, EA, MDD, and SCZ (Table 1) and 637 (p < 5e-8) SNP associations were found, including 322 variants that are reported for the first time for any trait (Supplementary Table 6) according to GWAS catalog. Two associations (rs2388334 and rs1452075) intersected with the twelve associations identified in the landmark common genetic variants of autism study (Grove et al. 2019). When assessed at the gene level, all 12 were concordant (as indicated in STable 6). Decomposition implemented in MetaPhat, using stepwise tracing of p-value and Bayesian information criteria (BIC) contributions (Lin et al. 2020; Schwarz 1978), identified 37 autism central trait SNPs where 16 were identified with multivariate GWAS approach (all relevant univariate SNPs p > 5e-8, listed in Supplementary Table 2). These 37 multivariate autism SNPs, 17 of which had previously been reported in existing GWAS studies, mapped to 35 genes (Table 2) and confirmed that 8/35 (ARHGAP32, CADPS, CUL3, KANSL1, MACROD2, MAPT, MSRA and NEGR1) are known curated SFARI genes, with autism susceptibility EAGLE scores <= 3 (indicating limited evidence) (Schaaf et al. 2020). The variant rs538628 within the NSF gene, a regulator of AMPA receptor endocytosis and critical for mediating glutamatergic synaptic transmission (Iwata et al. 2014), previously only implicated in mice with autism-like behaviors (Xie et al. 2021) and MAPT are identified to associate with the autism optimal central traits of autism, EA and SCZ (MAPT p = 3.98e-31, NSF p = 1.99e-27, Supplementary Table 2, trace plots are provided in supplementary data). Shown in the same table, MTAG (Turley et al. 2018) multivariate GWAS validation was performed to address potential cohort sample overlaps and similar results were found (MAPT p = 1.99e-20, NSF p = 5.37e-18).

**Table 2:** Multivariate GWAS autism-central SNPs tested with Coloc and MR tests towards the identified autism central traits, with all 8 traits passing MR and 19 gene regions/traits pairings passed Coloc (H4 > 0.90), indicated with +. Coloc and MR details are additionally listed in Supplementary Tables 3 and 4. The order of the central traits are determined by p-value importance during decomposition processing. Known GWAS associations (17/37) are marked as * while SFARI autism gene members (8) are in bold.

| rsid | Gene (SFARI Score) | Chr:pos:ref>alt | SNP consequence | Central traits |
| --- | --- | --- | --- | --- |
| rs6699841 | **NEGR1** | 1:72645850:A>G | intergenic | EA Autism |
| rs67980110 | ENSG00000237435 | 1:96470851:T>C | regulatory | EA Autism ADHD |
| rs2391769* | NA | 1:96978961:A>G | intergenic | Autism ADHD |
| rs58378462+ | ENSG00000221849 | 2:104138639:A>G | intron | Autism EA+ ADHDCHILD ADHD |
| rs11897599 | MRPS18BP2 | 2:140449566:A>G | downstream | EA Autism ADHD |
| rs78826721+ | NDUFS1 | 2:207002314:A>G | intron | SCZ+ EA Autism |
| rs6748341* | **CUL3** | 2:225377574:T>C | regulatory | Autism EA SCZ |
| rs1452075* | **CADPS** | 3:62481063:T>C | intron | EA Autism |
| rs62243489+ | **CADPS** | 3:62482927:T>G | intron | EA+ Autism |
| rs6806355+ | NA | 3:70488292:T>G | intergenic | BP SCZ+ EA+ Autism |
| rs35544582+ | SLC30A9 | 4:42044036:A>C | intron | EA+ Autism |
| rs67779882 | NA | 5:92488009:A>G | intron,non_coding | EA Autism ADHD |
| rs406413* | ENSG00000246316 | 5:113898581:T>C | intron | EA Autism |
| rs2388334* | ENSG00000271860 | 6:98591622:A>G | intron | BP EA Autism |
| rs6999466+* | **MSRA** | 8:10265712:A>G | intron | MDD EA+ Autism |
| rs877116* | ENSG00000253695 | 8:10712945:T>G | intron | Autism ASRD |
| rs2409743* | ENSG00000270076 | 8:11070360:T>G | intron,non_coding | Autism ASRD |
| rs2409784+ | BLK | 8:11396856:A>C | intergenic | Autism ASRD+ |
| rs11775333 | NA | 8:142637867:T>C | regulatory | EA Autism |
| rs11143599 | ENSG00000221844 | 9:76101777:T>G | intron,non_coding | BP Autism |
| rs1848797+* | ENSG00000238280 | 10:64552934:A>G | intron | BP+ EA Autism |
| rs12761761* | BNIP3 | 10:133775375:T>C | downstream | EA Autism |
| rs2237943 | SERGEF | 11:17838248:T>C | regulatory | EA Autism ADHD |
| rs4609618 | **ARHGAP32** | 11:128818792:A>C | intergenic | Autism ADHD |
| rs568828+ | NTM | 11:131732259:T>G | intergenic | Autism SCZ+ ADHD+ |
| rs177413+ | PSEN1 | 14:73683194:T>C | intergenic | BP+ Autism EA SCZ |
| rs736281* | NA | 14:94287830:T>C | intron | EA Autism |
| rs28929474* | SERPINA1 | 14:94844947:T>C | intron | EA Autism |
| rs62065453+* | ENSG00000131484 | 17:43573419:A>G | regulatory | Autism EA+ SCZ+ |
| rs62057107+* | CRHR1 | 17:43896032:T>C | intergenic | Autism EA+ SCZ+ |
| rs62061734+* | **MAPT** | 17:44018488:T>C | intron | Autism EA+ SCZ |
| rs2696633+ | **KANSL1** | 17:44270059:T>G | intron | Autism EA+ SCZ |
| rs538628* | NSF | 17:44787313:T>C | regulatory | Autism EA SCZ+ |
| rs1792709* | ENSG00000206129 | 18:53768975:A>G | intron | Autism SCZ |
| rs6079546 | **MACROD2** | 20:14716738:T>G | intergenic | MDD EA Autism |
| rs6035835 | XRN2 | 20:21271669:A>G | intergenic | Autism ADHDCHILD ADHD |
| rs9974470 | ENSG00000249209 | 21:35012066:A>G | intron | EA Autism |
| rs9974470 | ENSG00000249209 | 21:35012066:A>G | intron | EA Autism |

Shown in Supplementary Table 7, Figure 2e and Supplementary Figure 3, pathway enrichment using the 35 associated genes was performed with Enrichr (Kuleshov et al. 2016). Nervous systems development (GO:0007399) was found to be the most significant (p = 1.73e-08) while neural and microtubule structural related pathway hits from Reactome (Fabregat et al. 2018) and WikiPathways (Nesterova et al. 2019) featured pathways were Inclusion Body Myositis (MAPT and PSEN1, p = 1.27e-04) and COPII-mediated Vesicle Transport (NSF and SERPINA1, p = 4.69e-03) Additionally, human organ tissue system enrichment analysis was performed using WebCSEA (Dai et al. 2022) found significance with the digestive, nervous, sensory, lymphatic and respiratory organ systems (Figure 2f, p < 1e-03). As shown in Supplementary Figure 4, the most enriched tissue types are related to cerebrum, cortex, intestine and blood related components discerned from 1,355 tissue-type (TS) as well as data from the human brain single cell project (Lake et al. 2018).

**Figure 2:**
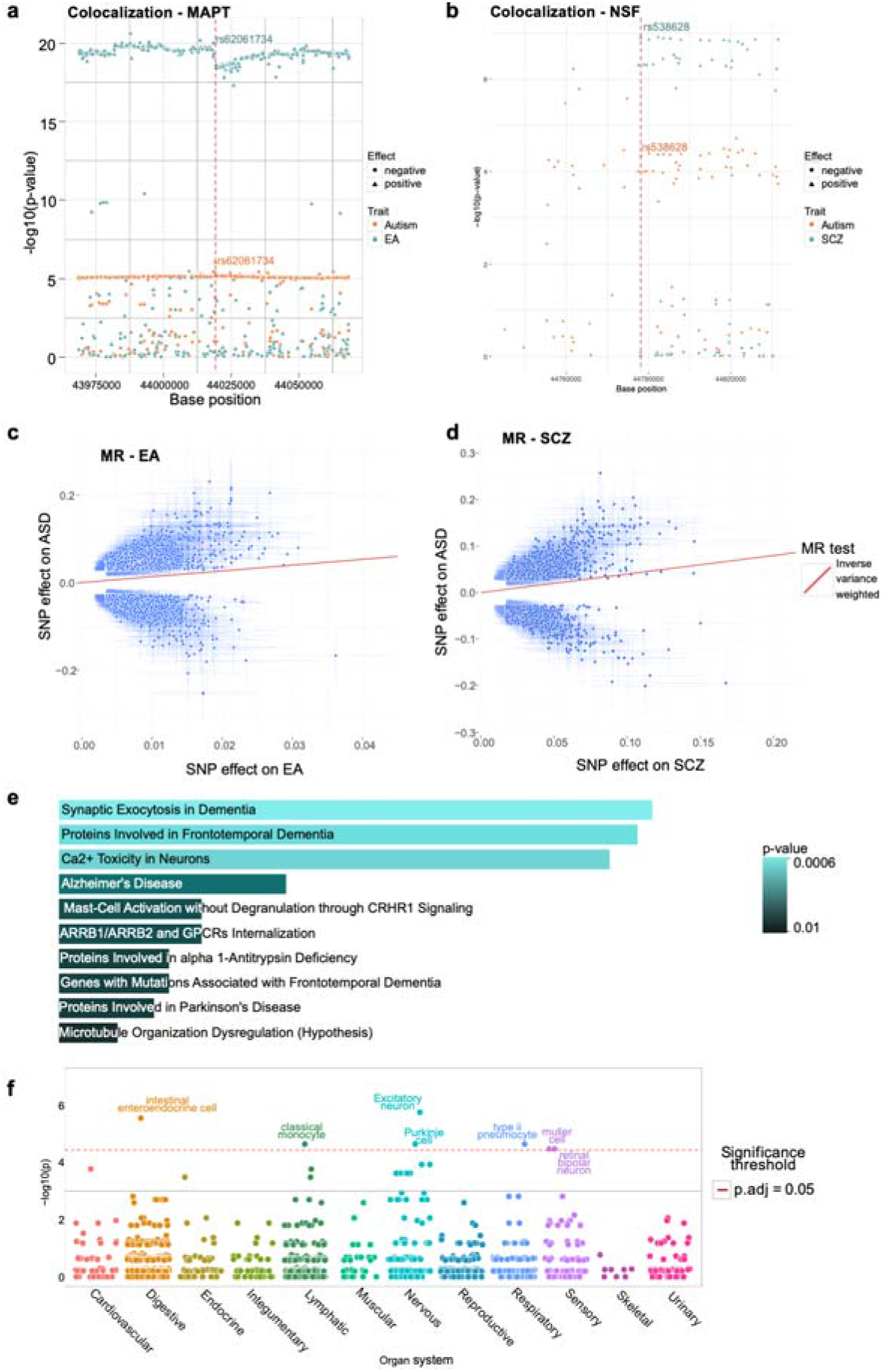
Results from the post GWAS analysis of the 37 selected SNPs. **a,b)** Colocalization processing using the original summary statistics of autism and EA for (a) rs62061734 (MAPT, failed colocalization with H4 probability 8.19%, p = 0.09), autism and NSF for (b) rs538628 (NSF, SCZ passed colocalization with H4 probability 0.94%, p = 1.1e-05), depicting supporting regional SNPs (x-axis) and their negative log10 p-value (y-axis) and effect direction (circles negative, triangles positive). **c,d)** Mendelian randomization (MR) results using inverse variance weighted (IVW) -method for association of autism SNP effects (y-axis) and c) EA and d) SCZ effects (x-axis). **e)** Pathway analysis for the genes associated with the selected SNPs show enrichment in processes related to neurons using Reactome database. The length of the bar represents the significance of that specific gene-set or pathway and the color indicates the significance of the pathway. Details of the pathways and genes with their associated p-values are listed in Supplementary Table 8. **f)** Organ system enrichment was applied using WebCSEA, using the selected 37 multivariate gene associations and found enrichment (p < 1e-03) with the autism relevant digestive, nervous and sensory organ systems as well as lymphatic and respiratory systems

### Colocalization analyses

Colocalization analysis was conducted on the 37 multivariate SNP associations identified to contain autism as a central trait. The comparative analysis was performed on the relevant mapped gene window, from start to end while adding 25 KBs on both ends to cover regulating and promoter regional elements. For the two SNPs that did not map to a gene, the window size used for the colocalization analysis was 100 KB (50 +-KB), estimated and derived from the gene median length of 24KB (Fuchs et al. 2014). Additional information concerning the number of regional LD adjusted SNPs applied to the colocalization test is shown in Supplementary Table 3. A total of 19/37 SNPs showed strong evidence for a common liability variant with autism (H4 > 0.9, details shown in Supplementary Table 3) and the traits having common autism liable variants included EA (9), SCZ (6), BP (2), ADHD (1) and ASRD (1). Notably. SNP rs62061734, mapping to the MAPT gene and rs538628, mapping to the NSF gene had H4 of 99% for EA and SCZ, respectively (shown in Figure 2a-b) while SNP rs568828, mapping to the NTM gene had H4 of 99% for SCZ and ADHD (Supplementary Table 2).

### Mendelian randomization analyses

Mendelian randomization analysis was conducted for the 8 central traits genetically correlated with autism. The lead SNPs, with F1 scores > 25 (listed in Supplementary Table 4, where > 10 is considered strong (Palmer et al. 2012)) were found to lend significantly increase probability of autism (p < 0.001 both IVW and MR-Egger (EA and SCZ are shown in Figure 2c-d), accounting for horizontal pleiotropy and multiple testing with Bonferroni correction of 8 traits), after checking for directionality with MR Steiger testing.

### Validation

The associations of MAPT and NSF genes are among the affected autism hits in the concordant SNPs. To assess the impact of the reported multivariate associations on expression (eQTL) and splicing regulatory quantitative trait loci across tissues, the majority (22/37 eQTL, 24/37 sQTL, details listed in Supplementary Table 9) of the associations found are cited in the EBI QTL Catalog (Kerimov et al. 2021) where they associate (adjusted p < .05) with adipose, brain and neuron tissues. Furthermore, filtering on GeneCards (Safran et al. 2010) curations, the presented autism central genes are enriched with systems related to gut, microbiome, intestinal immune, enteric nervous and central nervous systems (Supplementary Table 5).

Additionally, the distribution of these autism-central trait related SNPs in 97 (30 autism proband) family based GEMMA (Troisi et al. 2020) samples was investigated. Visualized in Figure 3, distinct clustering and proportional differences between the 30 proband and 67 controls were found with repect to the neurological developmental genes MAPT and NSF were found. MAPT SNP was present in 7/30 (23%) probands compared to 25/67 (37%) controls while for NSF was enriched in 24/67 of the controls as compared to 4/30 (13%) probands. Shown in Supplemntary Table 8, the phi coefficients for MAPT, NSF and KANSL SNPs between probands and controls were > 0.2, indicating strong correlations. Separated by approximately 770K bases apart, these two genes are both found in the 17q21.31 arm and referred to as the MAPT H1/H2 haplotypes (Supplementary Table 8). This haplotype, associated with the Tau microtubule-associated protein, has been previously implicated in Parkinson’s disease and neurological development disorders (Wang and Mandelkow 2016; Wider et al. 2010). After overlaying the autism multivariate-GWAS variants in the context of the Grove et al. (Grove et al. 2019) study as a Manhattan plot (Supplementary Figure 5), capturing KANSL1, MAPT and NSF, the reported MAPT and NSF autism linked SNPs were found to be enriched with the MAPT H2 haplotype (shown in Figure 3), as well as a deletion event located in intron 9 of MAPT, thus impacting MAPT alternate splicing and potentially reduction aggregation of Tau (Corsi et al. 2022).

**Figure 3:**
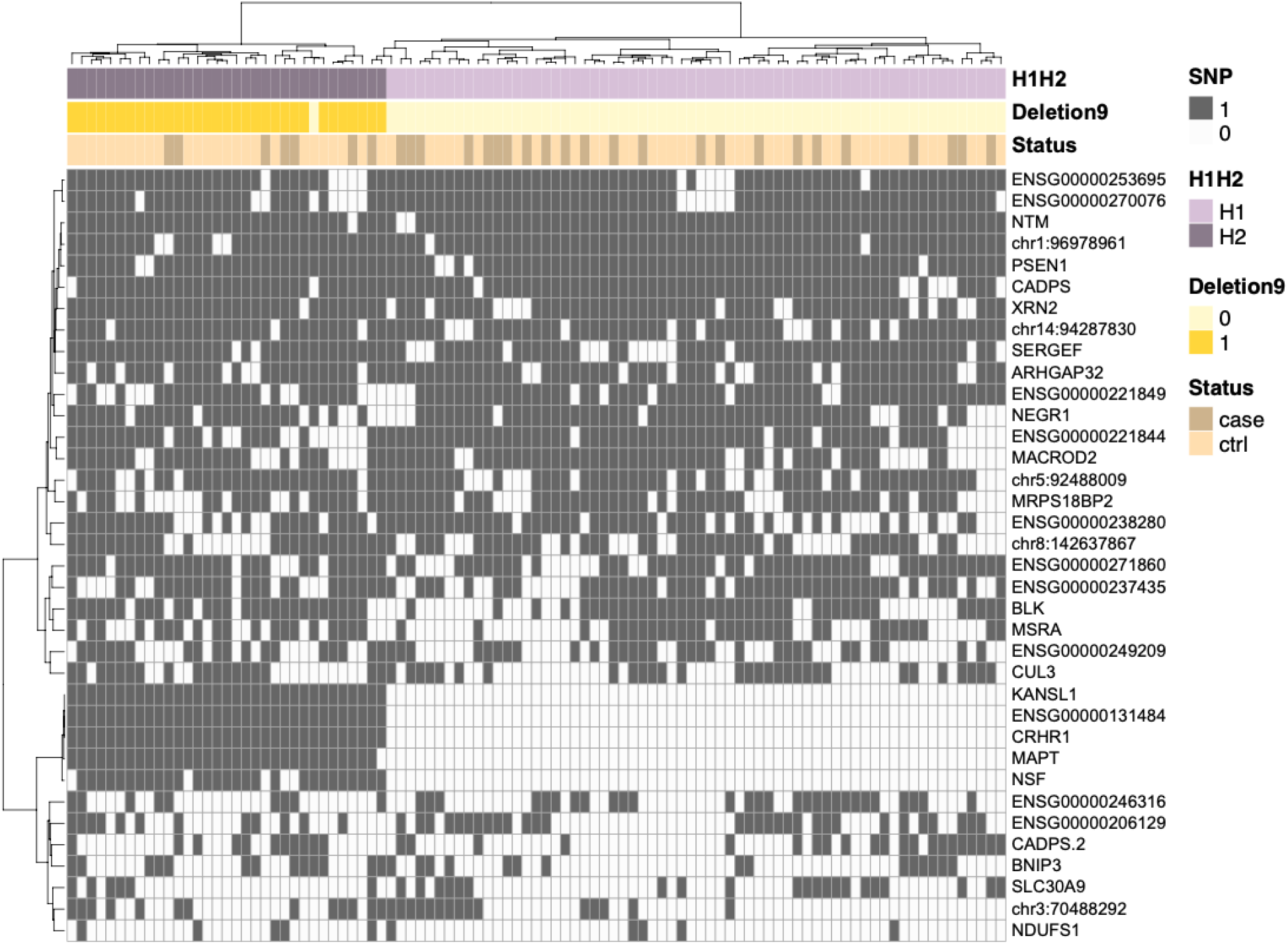
Results from the validation with GEMMA -cohort. Heatmap shows the clustering of the selected SNPs in the cohort, with rows showing the genes associated to each SNP and samples shown in columns. Samples with structural variation in chr17q21.31 can be seen to cluster together on the left. Darker yellow annotation highlights the samples with the 238bp deletion located in intron 9 of MAPT -gene, also associated with the H2 haplotype. Assessment of the H1/H2 haplotype for MAPT was evaluated using SNPs rs1052553 and rs8070723. H1 haplotype has been shown to increase risk of other neurological disorders such as Altzheimer and Parkinson’s Disease, and as seen in the GEMMA cohort, autism probands tend to express the H1 haplotype in lighter purple (23 out of 30).

## Discussion

Using multivariate statistical learning approaches, this study constitutes the largest and most comprehensive genetically correlated multi-trait GWAS analysis with summary statistics performed on autism and its genetically correlated traits; ADHD, ADHD childhood, ASRD, bipolar, DBD, EA, MDD, and SCZ to explore the underpinnings driving the complexities in autism. 37 associations containing autism as a central trait were discovered, with 16 of these associations were detected only due to the increased statistical power of this multivariate GWAS analysis (lowest univariate p-value all traits > 5e-08, and 12/16 confirmed with the MTAG tool (Turley et al. 2018), Supplementary Table 2). Enrichment analysis confirmed that the multivariate autism association results are related to neuron and gut tissues and developmental pathways as well as inflammation and microbiome domains, further underscoring the intersection of genome and microbiome as well as supportive of the gut-brain axis hypothesis associated to autism (Morton et al. 2023; Cheng et al. 2023). The genetic associations pertaining to MAPT, NSF and NTM are known markers to microtubule dysregulation and cytoskeleton pathways (Supplementary Table 7) and add additional support to the canon of impairments in the regulation of the brain-barrier and gut permeability underpinning autism development (Fiorentino et al. 2016). Using the multivariate autism central trait gene sets, based on comprehensive human tissue cell type and single cell data (Dai et al. 2022, Lake et al. 2018) analysis, enrichments were detected with digestive, nervous, and sensory organ systems (Figure 2f). At the tissue cell type level and further supporting the gut-brain axis and blood brain barrier, the analysis detected enriched autism relevant signals related to brain, adipose and gut eQTL/sQTL (Supplementary Table 9) tissue panels.

Overall, the identified autism biomarkers passed MR with strong F1 measures and significantly contributed to improve the construction of meta psychiatric based autism polygenic scores (Jansen et al. 2020), shown to improve prediction relative to standard PRS in other complex conditions such as coronary heart disease and type 2 diabetes (Lin et al. 2023; Tamlander et al. 2022). These multivariate autism annotations are mapped to genes including MAPT and NSF that are known members of biological pathways driving neural disorders such as infantile epilepsy (Suzuki et al. 2019) and Parkinson’s Disease (Brion, Octave, and Couck 1994; Derkinderen et al. 2021). Interestingly, colocalization tests for the MAPT region indicated shared genetic risk between only EA and autism (H4 0.99), while that for the NSF gene did not associate with EA, instead associated with SCZ (H4 0.94), suggesting intra region heterogeneity that demands future investigation. A MAPT association (rs17649553) has recently been cited as a strong (p = 4.86×10-37) association with protective effect towards Parkinson’s Disease (Nalls et al. 2014) as well as liable eQTL towards autism (Dominguez-Alonso, Carracedo, and Rodriguez-Fontenla 2023). With respect to autism, the KANSL1, BNIP3, CADPS and NEGR1 genes have been implicated with immune and microbiome features (Cheng et al. 2023) and behavioral developments (Singh et al. 2019).

The most common traits in our set of 37 associations that passed colocalization with autism were EA (9), SCZ (6) and BP (2). It is known that the diagnosis for autism and ADHD, particularly ADHD manifestation in young children, is similar with symptomatic issues concerning hyperactivity and attention span (Kern et al. 2015). While a previous study has performed comparison of genetic and functional enrichment of associations between autism and ADHD (Peyre et al. 2021) GWAS resources, this study further complements their results by inclusion of other autism co-occurring traits, including ADHD and ADHD childhood as well as EA. Interestingly, autism and ADHD have both been linked with dysbiosis disruption in microbiome composition and function, gastrointestinal and bowel habits issues (Morton et al. 2023).

As part of validation, clustering and distribution proportion differences based on the autism identified SNP associations were detected between probands and non-autistic subjects on genomes from the GEMMA cohort (Troisi et al. 2020). The MAPT haplotype region (17q21.31) has been linked with neurological disorder development and general cognitive functions (Trampush et al. 2017), here intron 9 deletion associated to structural aberration event was highly depleted in autism probands (23%, 7/30). Deletion and its associated inversion event are known to disrupt alternate splicing and decrease the aggregation of tau, which would lead to instability and structural impairment in the brain neuronal cells (Avila et al. 2004). Our validation results are limited by the relatively small sample size (97) currently available in GEMMA. Nevertheless, the independent and deep sequencing data has allowed the harvesting of interesting observations concerning distribution autism-central trait associations in probands as compared to familial controls. The future release of other omics data types from autism cohorts and foundation, including additional microbiome, metabolome and methylation measurements, will allow for improved statistical power with deeper temporal analysis towards confirmation of gut-brain-axis changes and genetic patterns driving autism heterogeneity and development.

## Conclusion

Our study represents the largest autism multi-trait GWAS, analysis conducted to date, combining autism and 8 related trait genome-wide association summaries and performed systematic decomposition to identify novel genetic associations related to autism and autism co-occurring driver traits. Upon analysis with colocalization and Mendelian randomisation, EA, SCZ, BP and ADHD associations were found to share genetic risks with autism and potentially liable, signaling enrichment of potentially liable patterns within brain tissues and cell types implicated with neurodevelopment and the gut-brain axis hypothesis.

## Data Availability

All suumary data produced in the present study are available upon reasonable request to the authors

## Acknowledgements

The work was supported by the European Commission Horizon 2020 programme (grant no. 825033). We are sincerely grateful to the entire GEMMA team and in particular to the families participating in the project.

## Conflicts of Interest

The authors have no conflicts of interests.

## Author contributions

JL, KS, CLA, CR, MN and RA designed the study. NV, KS, ST, RA and JL performed the data analysis and integration. KS, NV, ST, FI, RA and JL conducted the validation and figure generation. KS, NV, AF, CR, RA and JL wrote the paper. All authors contributed to critical revisions and approved the final manuscript.

## Supplementary Figure Legends

**Supplementary Figure 1:** Genetic correlation of the traits included in the analysis. ASD=Autism Spectrum Disorder; ADHD=Attention Deficit Hyper Disorder; ASRD=Anxiety-Stress Disorder; DBD=Disruptive Behaviour Disorder; EA=Education attainment; MDD=Major Depression Disorder; SCZ=Schizophrenia.

**Supplementary Figure 2:** P-value and BIC decomposition processing of MAPT and NSF to identify autism central traits. ASD=Autism Spectrum Disorder; ADHD=Attention Deficit Hyper Disorder; ASRD=Anxiety-Stress Disorder; DBD=Disruptive Behaviour Disorder; EA=Education attainment; MDD=Major Depression Disorder; SCZ=Schizophrenia.

**Supplementary Figure 3:** Pathway analysis using the WikiPathway database also highlights neuronal processes, with bar length and color indicating significance. More details listed in Supplementary Table 8.

**Supplementary Figure 4:** Tissue and cell (TS) type enrichment using WebCSEA and the list of the 22 central trait genes found that the most enriched tissues are related to cerebrum, cortex and small intestine related tissue types. Lake 2017 refers to data from human brain single cell analysis project (https://pubmed.ncbi.nlm.nih.gov/29227469/) while HCA stands for histologic chorioamnionitis, an intrauterine inflammatory condition.

**Supplementary Figure 5:** Autism multivariate GWAS associations within the MAPT H1/H2 haplotype, 17q21 arm region, are presented in a Manhattan plot, in the context of Grove et al. GWAS results. Significance thresholds for p-values of 1e-05 indicated in blue and 1e-08 in red. Significant SNPs highlighted in green show rs62061734 (MAPT), rs269633 (KANSL1) and rs538628 (NSF).

## Supplementary Tables

Supplementary Table 1: Data and sample details of autism and 8 genetically correlated traits (P < 0.05, calculated from LDSC) are presented and applied towards multivariate-GWAS. Data from four excluded traits are additionally shown.

Supplementary Table 2: 37 multivariate associations are identified with autism as a central trait where 17/37, shown with asterisk are previously reported in the GWAS Catalog and in bold, 8 genes are identified as SFARI autism genes.

SupplementaRY Table 3: 19 gene regions/trait pairings passed coloc (Posterior Prob H4 > 0.9, Shown in bold,) called on coloc.abf with a window size of ± 50 kb flanking the SNP locus

Supplementary Table 4:(A) Mendelian randomization (MR) results for autism as outcome and related traits. (B) MR where autism is the exposure and related traits are the outcome.

Supplementary Table 5: MV associated genes are found in systems curated/implicated with gut microbiome and neural systems from GeneCards.

Supplementary Table 6: List of 637 Significant SNPs (p < 5e-8), with 315 already reported in the GWAS catalog, identified by MetaPhat multivariate-GWAS using autism and 8 genetically correlated trait summary statistics.

Supplementary Table 7: A) 108 enriched (p < 0.05) Go terms are annotated and (B) 46 pathways on WikiPathway C) KEGG D) Reactome resources e) Tissue from the list of multivariate autism SNPs found enrichments in neuron and nervous systems related data.

Supplementary Table 8: Autism central SNP alleles are mapped to GEMMA genotypes called from WGS.

Supplementary Table 9: eQTL and sQTL related results of the autism central associations relative to brain and nervous systems from EBI QTL catalog are captured via https://fivex.sph.umich.edu/. Study URLs are listed at the bottom of the table.

## Notes

### Competing Interest Statement

The authors have declared no competing interest.

### Funding Statement

The work was supported by European Commission Horizon 2020 programme (grant no. 825033)

### Summary of Updates

LDSC was employed to select and filter ASD related traits based on genetic correlation. MTAG was applied to validate multivariate GWAS signals yielded from MetaPhat/MetaCCA. Selected associations were further assessed using QTL. Larger sample sizes from GEMMA were integrated into the validation step.

