## Supplemental Figures for "Exploring autism and autism co-occurring condition associations to elucidate multivariate genetic mechanisms and insights"

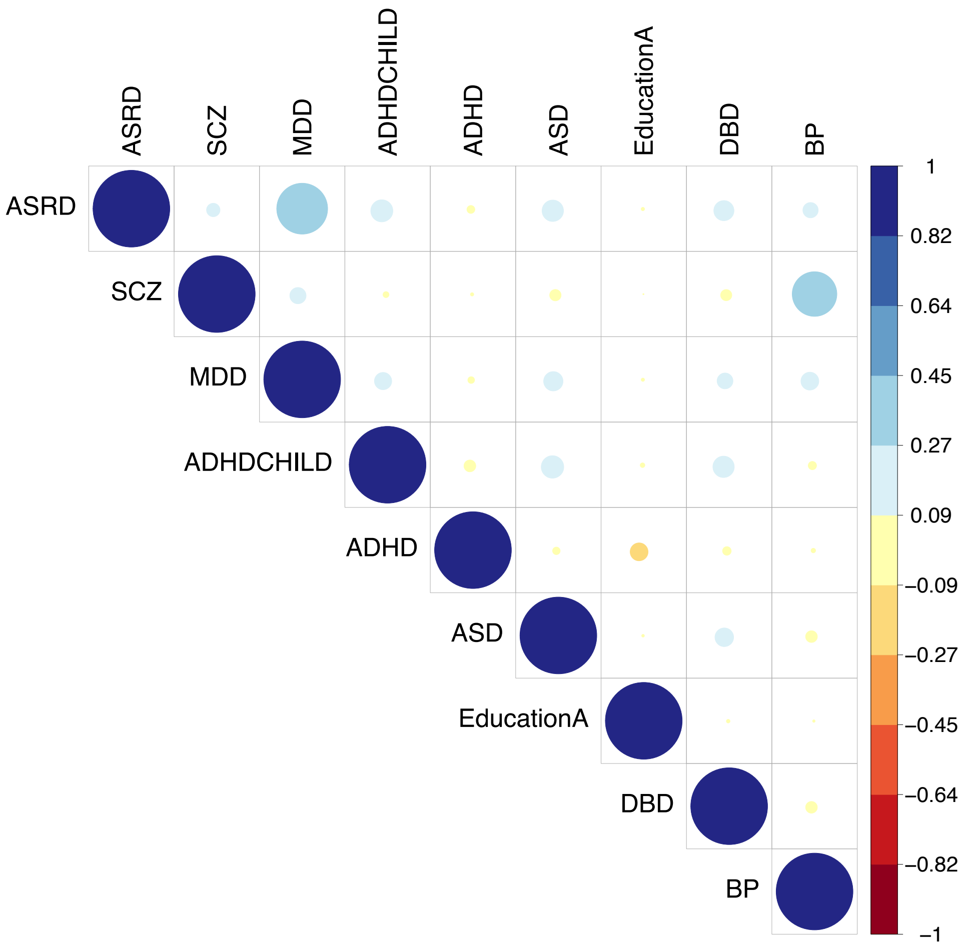


*Supplementary Figure 1*: Genetic correlation of the traits included in the analysis. ASD=Autism Spectrum Disorder; ADHD=Attention Deficit Hyper Disorder; ASRD=Anxiety-Stress Disorder; DBD=Disruptive Behaviour Disorder; EA=Education attainment; MDD=Major Depression Disorder; SCZ=Schizophrenia.

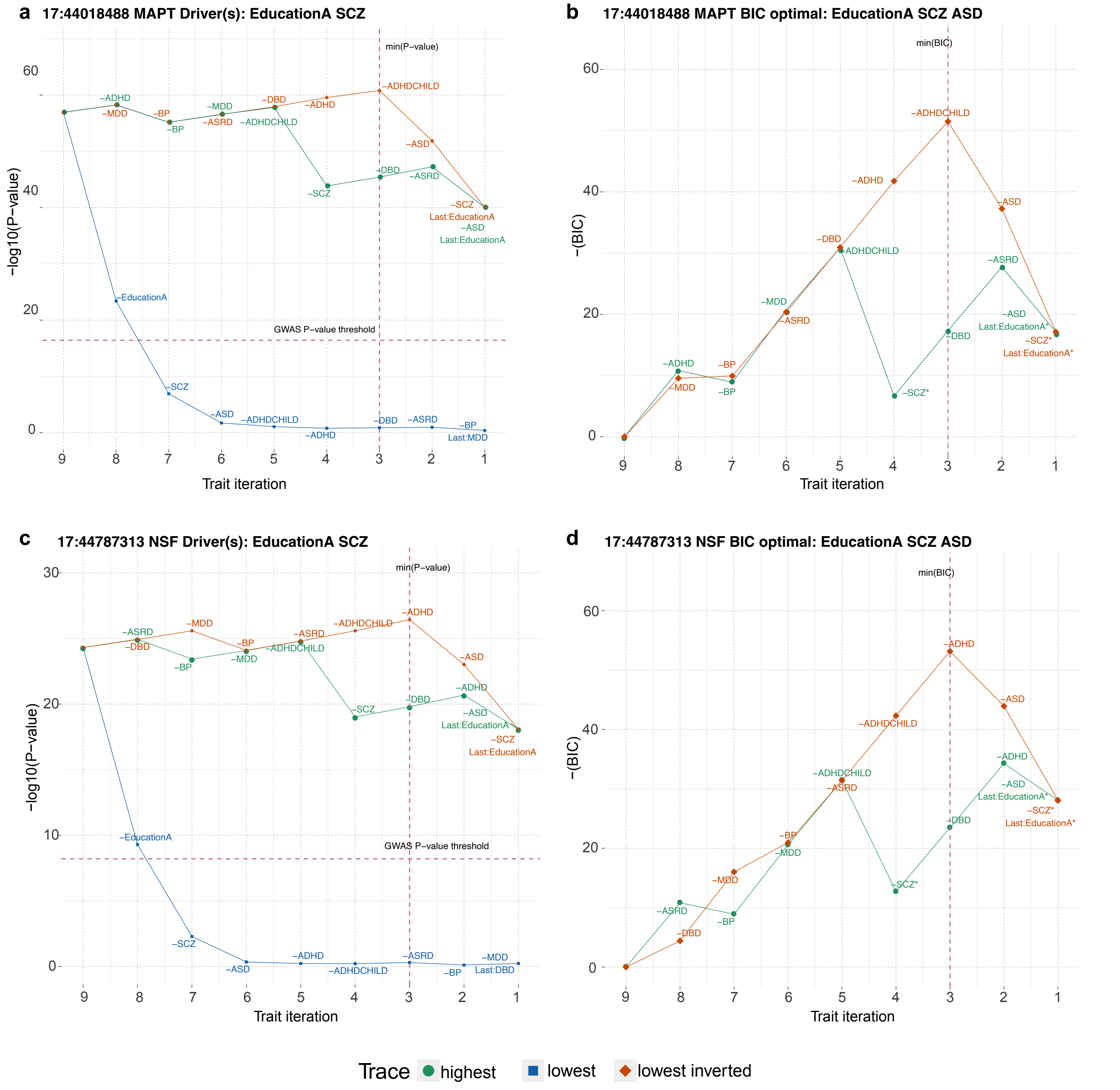


*Supplementary Figure 2:* P-value and BIC decomposition processing of MAPT and NSF to identify autism central traits. ASD=Autism Spectrum Disorder; ADHD=Attention Deficit Hyper Disorder; ASRD=Anxiety-Stress Disorder; DBD=Disruptive Behaviour Disorder; EA=Education attainment; MDD=Major Depression Disorder; SCZ=Schizophrenia.

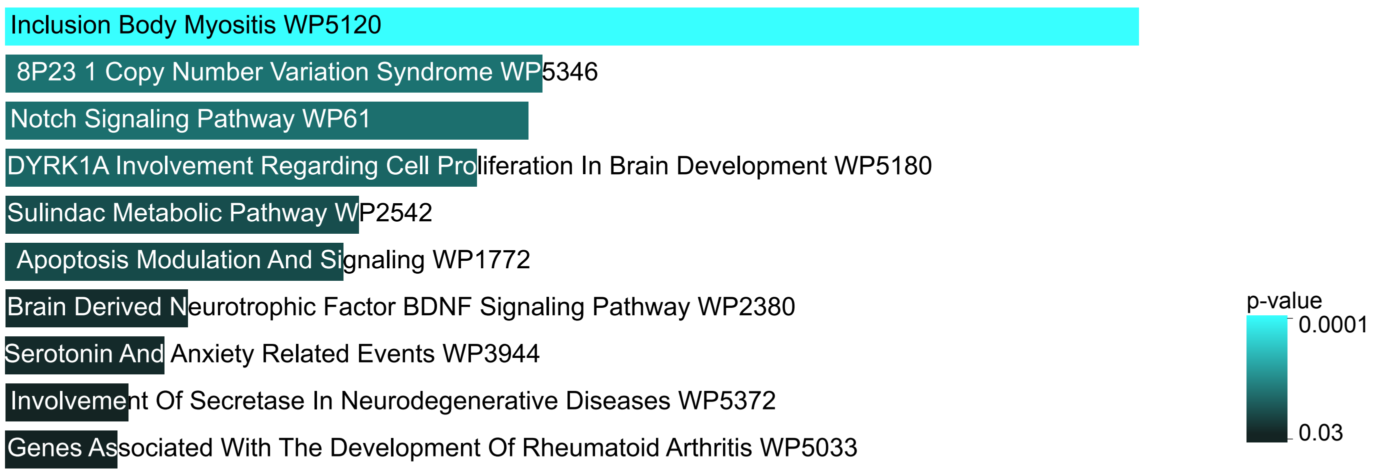


*Supplementary Figure 3:* Pathway analysis using the WikiPathway database also highlights neuronal processes, with bar length and color indicating significance. More details of enriched pathways listed in Supplementary Table 8.

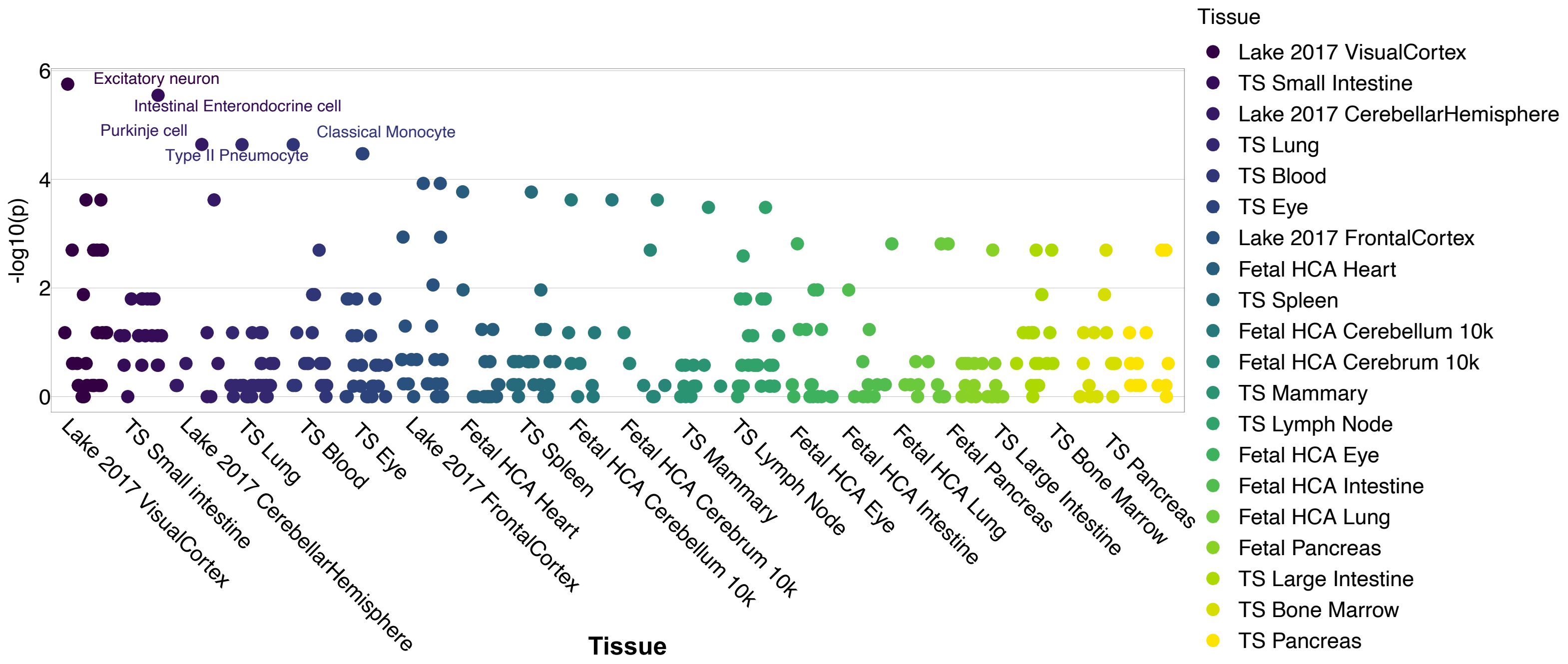


*Supplementary Figure 4:* Tissue and cell (TS) type enrichment using WebCSEA and the list of the 22 central trait genes found that the most enriched tissues are related to cerebrum, cortex and intestine related tissue types. Lake 2017 refers to data from human brain single cell analysis project (https://pubmed.ncbi.nlm.nih.gov/29227469/) while HCA stands for histologic chorioamnionitis, an intrauterine inflammatory condition.

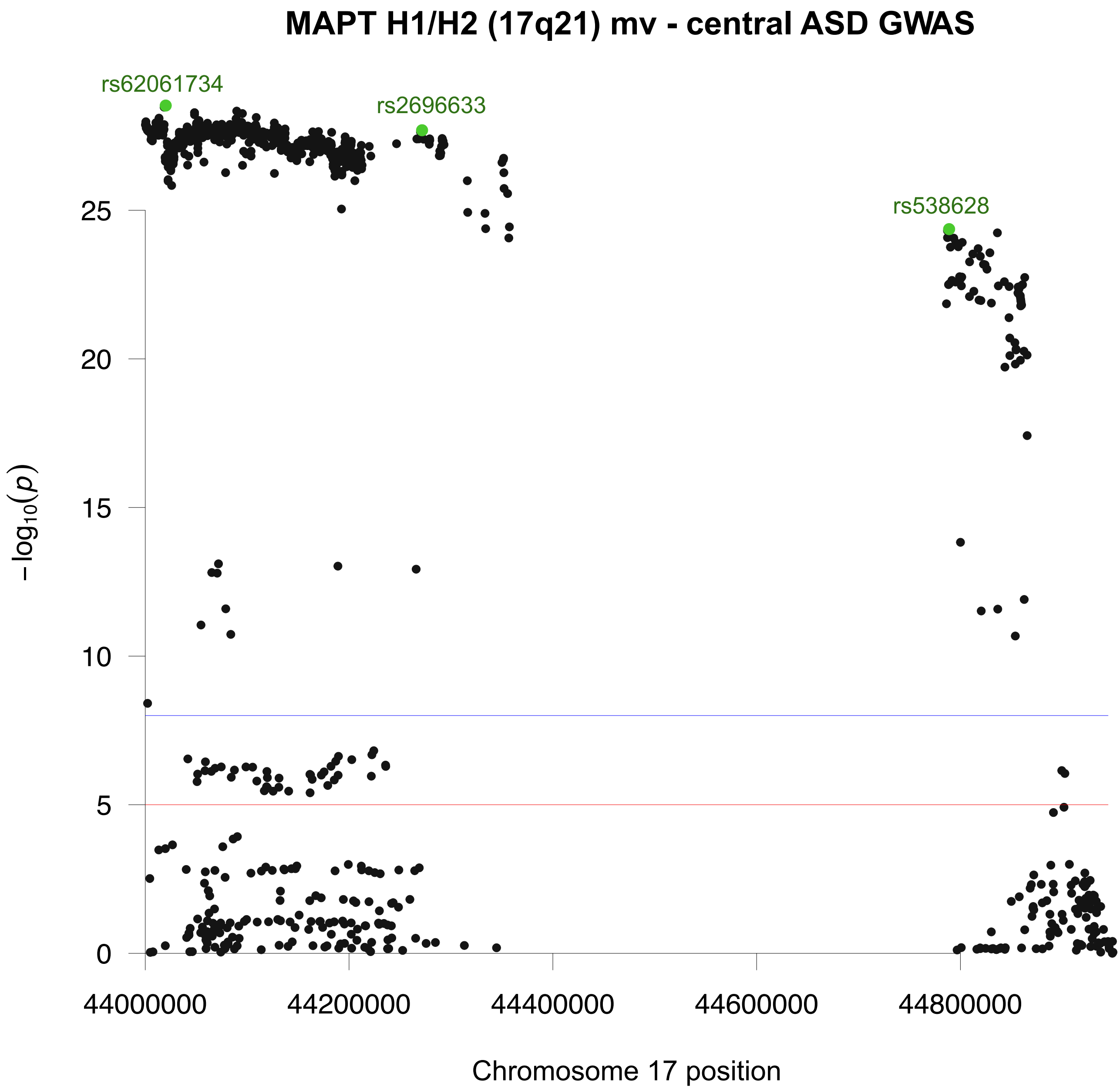


*Supplementary Figure 5:* Autism multivariate GWAS associations within the MAPT H1/H2 haplotype, 17q21 arm region, are presented in a Manhattan plot, in the context of Grove et al. GWAS results. Significance thresholds for p-values of 1e-05 indicated in blue and 1e-08 in red. Significant SNPs highlighted in green show rs62061734 (MAPT), rs269633 (KANSL1) and rs538628 (NSF).
